# Patient attrition in Molecular Tumour Boards: A Review

**DOI:** 10.1101/2021.10.07.21264241

**Authors:** Hannah Frost, Donna M. Graham, Louise Carter, Paul O’Regan, Donal Landers, Andre Freitas

## Abstract

Molecular Tumour Boards (MTBs) were created with the purpose of supporting clinical decision making within precision medicine. Though these meetings are in use globally reporting often focuses on the small percentages of patients that receive treatment via this process and are less likely to report on, and assess, patients who do not receive treatment. A literature review was performed to understand patient attrition within MTBs and barriers to patients receiving treatment. A total of 54 papers were reviewed spanning a 6 year period from 11 different countries. 20% of patients received treatment through the MTB process. Of those that did not receive treatment the main reasons were no mutations identified (26%), no actionable mutations (22%) and clinical deterioration (15%). However, the data was often incomplete due to inconsistent reporting of MTBs with only 53% reporting on patients having no mutations, 48% reporting on presence of actionable mutations with no treatment options and 57% reporting on clinical deterioration. As patient attrition in MTBs is an issue which is very rarely alluded to in reporting, more transparent reporting is needed to understand barriers to treatment and integration of new technologies is required to process increasing omic and treatment data.

## Introduction

The human genome project provided the world with a fully referenced genome that helped to illuminate the role of somatic and germline mutations in the pathogenesis of cancer^1^. The development of next generation sequencing (NGS) propelled genomics research even further, enabling sequencing of entire genomes within days rather than decades. This helped facilitate the use of genomics sequencing within clinically meaningful timelines and identify aberrant pathways for the development of new and effective targeted treatment options for patients^2^, facilitating rapid precision medicine on a larger scale^3^. Precision medicine is a healthcare model that allows treatment to be tailored to individuals by categorising them into subpopulations^4^. Precision medicine is reliant on the knowledge and expertise of coordinating specialities, requiring persistent adoption of rapidly advancing science and new techniques.

Targeted therapies are drugs that target specific genes or proteins in cancerous cells^5^. Lung cancer treatment has had a number of successes with targeted therapy; drugs targeting EGFR mutations and ALK and ROS1 translocations are now routinely used in cancer treatment^6^. However, precision medicine successes aren’t straightforward, for example, before the MET inhibitor Crizotinib was licenced for use in ALK translocations, it was originally tested in MET mutated tumours^7^. Despite having potent activity against MET^8^, studies found no anti-tumour effect in tumours with MET mutations^9^. Often determining appropriate targeted therapies for patients with pathogenic mutations requires input from multiple disciplines, therefore, organisations regularly consult with or develop Molecular Tumour Boards (MTB). Molecular Tumour Boards, Precision Genomics Boards or Genomics Review Boards are all names for a multidisciplinary team that consult on individual patients’ treatment options either providing expert opinion to healthcare professionals who have limited access to multidisciplinary expertise or driving decisions for their own patients. These meetings focus on patients with rare, hard-to-treat or late stage malignant disease and are composed of various specialists but always include oncologists or clinicians and scientists or biologists^10,11,20–29,12,30–39,13,40–49,14,50–59,15,60–63,16–19^. Clinical research has shown that these teams can help facilitate precision medicine, however, with increased evaluation limitations have emerged^64,65^.

MTBs were developed for the specific purpose of supporting complex clinical decision making and are often only reported in terms of positive outcomes. However, it is imperative that we are cognisant of the outcomes of patients who never reach the treatment phase on these pathways, to ensure that we are striving to improve processes and therefore opportunities for patients. This review aims to assess how global MTBs are conducted and identify common reasons for lack of treatment options, evaluating whether there are procedural issues that contribute to this attrition and areas for potential process optimisation. Additionally, suggested guidelines for the future reporting of MTBs may be informed by this review.

These guidelines could allow for transparent and consistent reporting, bringing awareness to deficiencies in the current system and facilitating change to mitigate against attrition and to ensure that all patients are given the greatest opportunity to access treatments.

The contribution of this paper is:

- Quantification of the issues with MTBs.
- A description of the reasons for patient attrition in an MTB.
- Recommendations for guidelines for optimal reporting of MTBs.

## Methodology

### Literature Based Analysis

A review of published literature was performed to evaluate current MTB processes and understand the reasons a treatment option is not identified or accessed by a patient after review in an MTB. Databases searched were EMBASE and PubMed, and last accessed 19 November 2020. Search terms were formalised for reproducibility purposes.

Inclusion criteria were:

- written in English.
- more than five patients were reviewed through the MTB
- multi-gene panel
- humans only
- and MTBs were either self-identified by the authors of the paper or were defined as a multidisciplinary team meeting that performed and reviewed multi-omic testing outside of standard of care, on patients with cancer, with an aim to finding a targeted therapy.

Exclusion criteria were:

- studies earlier than 2014
- non-oncology studies
- case studies
- imaging studies
- biomarker reviews
- evaluated specific treatment regimens or focused on specific mutations only
- an abstract, unless it supported a full paper
- had no centralised review of patients e.g., MTB
- no intention to treat
- or reported only in percentages making total numbers impossible to determine.

The country the MTB was based was recorded as well as the type of institution the MTB was held at, eligible cancer types, duration of the MTB, method of genomic testing performed, variant allele fraction threshold for action, and reasoning why patients were unable to access treatments.

Where numbers were unclear or reasons for attrition were grouped these were excluded from the analysis.

### Process Flow

Using the papers selected for review a systematic formalisation of the MTB process was created using papers which described their MTB patient pathway. At the end of the review all process flows were assimilated to create a universal structure. The flows included the patient journey from consent to tissue acquisition and analysis, the point at which patients were discussed at an MTB, the return of their full genomic results and how the results were disseminated.

The following categories for attrition were identified from the review: insufficient tissue; no mutations identified; no actionable mutations identified; actionable mutations identified but no treatment available; actionable mutations identified by ineligible for treatment; patient had already received the matched drug; off licence treatment available but couldn’t access; clinically deteriorated and patients were categorised accordingly. As each study did not report on all these categories there were multiple missing data points.

### Statistical analysis

Descriptive statistics only are provided due to the number of missing data points a more formal statistical review was deemed inappropriate. Patient attrition is described using percentages, as all papers did not report on each reason for attrition the percentages were derived only from papers that reported on them.

## Results

A literature review gave a fuller understanding of the global picture for patients and provided insight on the perceived importance by researchers of patient attrition. The review produced over 8000 targeted results (EMBASE 115/ PubMed 7888) which was reduced to 54 evaluable papers using the exclusion and inclusion criteria listed in the methods. All reviews and data collection were performed by a single reviewer.

### Study Characteristics

A summary of MTB characteristics can be found in Table 1. Exactly half of all studies enrolled 100 patients or fewer to an MTB, with an overall range of 14–3737. The average study length was 29 months (range 6-60 months).

**Table 1.** Search terms used in this review for both PubMed and EMBASE.

| Database | Search Terms |
| --- | --- |
| PubMed | **(((((study).ti,ab OR (trial).ti,ab) AND (review).ti,ab) AND ((cancer).ti,ab OR (oncology).ti,ab OR (tumour).ti,ab)) AND ((precision medicine).ti,ab OR (molecular tumour board).ti,ab OR (Institutional Review Board).ti,ab)) AND ((genomic profiling).ti,ab OR (precision oncology).ti,ab))** |
| EMBASE | **"((((genomic profiling).ti,ab OR "PERSONALIZED MEDICINE"/ OR (precision medicine).ti,ab OR (molecular tumour board).ti,ab OR (precision oncology).ti,ab) AND ("STUDY, PILOT"/ OR "STUDY, SINGLE BLIND"/ OR "STUDY,MULTICENTER"/ OR "STUDY,PROSPECTIVE"/ OR (study).ti,ab OR (trial).ti,ab OR "CLINICAL TRIAL"/ OR "ADAPTIVE CLINICAL TRIAL"/ OR "CONTROLLED CLINICAL TRIAL"/ OR "MULTICENTER STUDY"/ OR "PHASE 1 CLINICAL TRIAL"/ OR "PHASE 2 CLINICAL TRIAL"/ OR "PHASE 3 CLINICAL TRIAL"/ OR "PHASE 4 CLINICAL TRIAL"/ OR "CLINICAL TRIAL (TOPIC)"/)) AND (NEOPLASM/ OR "MALIGNANT NEOPLASM"/ OR "ADVANCED CANCER"/ OR "CHILDHOOD CANCER"/ OR "MULTIPLE CANCER"/ OR "PRIMARY TUMOR"/ OR "SECOND CANCER"/ OR "SOLID MALIGNANT NEOPLASM"/ OR "MALIGNANT NEOPLASM,SOLID"/ OR "MALIGNANT NEOPLASTIC DISEASE"/ OR (cancer).ti,ab OR (tumour).ti,ab OR ONCOLOGY/)) [DT 2020-2015] [Publication types Article OR Conference Abstract OR Conference Paper OR Conference Proceeding OR Conference Review OR Editorial OR Erratum OR Journal OR Report OR Review OR Short Survey OR Trade Journal] [English language] [Languages English] [Humans]"** |

### Process Flow

The overarching process flow was used to identify steps in the process where there is typically patient attrition (figure 1). Few MTBs had unique processes, those that did differ varied by bioinformatic pipeline and whether patients were presented to the MTB before and after profiling or after only. Other areas where MTBs differed were in how the results were disseminated. This was typically done in one or more of the following routes; through an online database; via patient health records; through email or phone call to the patient; or within a report given to the treating clinician.

**Figure 1.**
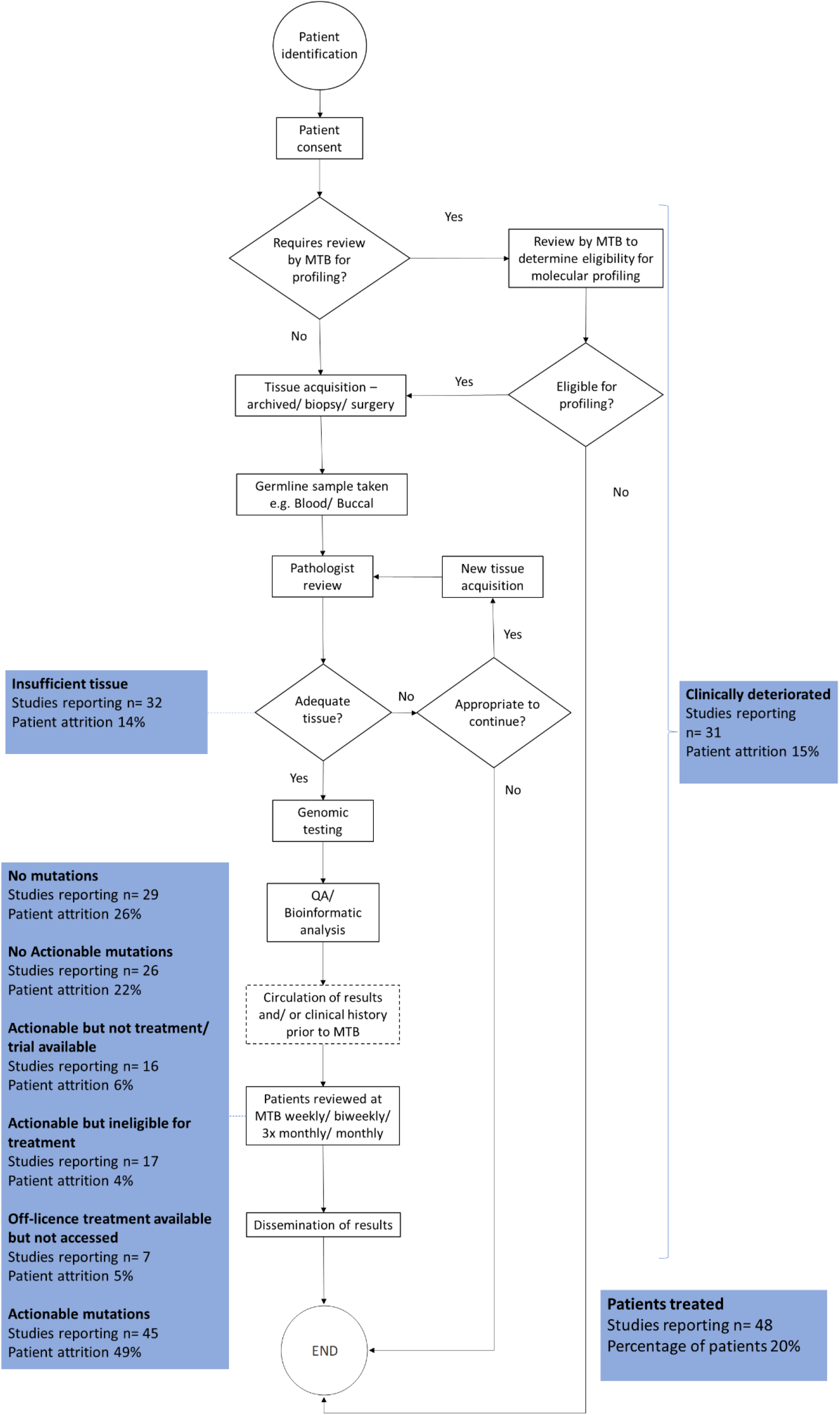
Process flow for MTBs globally with common reasons for attrition. As not all studies reported on all the reasons outlined in this review percentages were calculated out of the studies where the data was available. Abbrevations: MTB Molecular tumour board, QA Quality assurance.

### Patient Attrition

A total of 19686 cases were described within the selected papers. Of these the majority were adults (n= 16427), with 628 childhood cancer cases reported. 2631 cases were reported in a mixed adult and paediatric MTB. The reasons for not receiving therapy were inconsistently reported in the published literature and patient numbers reduced without explanation (unknown outcome n=5725, see Figure 2), therefore there is variability in data available for patient outcomes. Of those cases where the outcome was known, the most common reasons reported for patient attrition were: no mutations detected (26%), no actionable mutations detected (22%), clinical deterioration (15%) or lack of tissue (13%). The reason for the greatest number of patients not receiving treatment in paediatric trials (14%) was no actionable mutations, whereas in adult trials these were no mutations and no actionable mutations.

**Figure 2.**
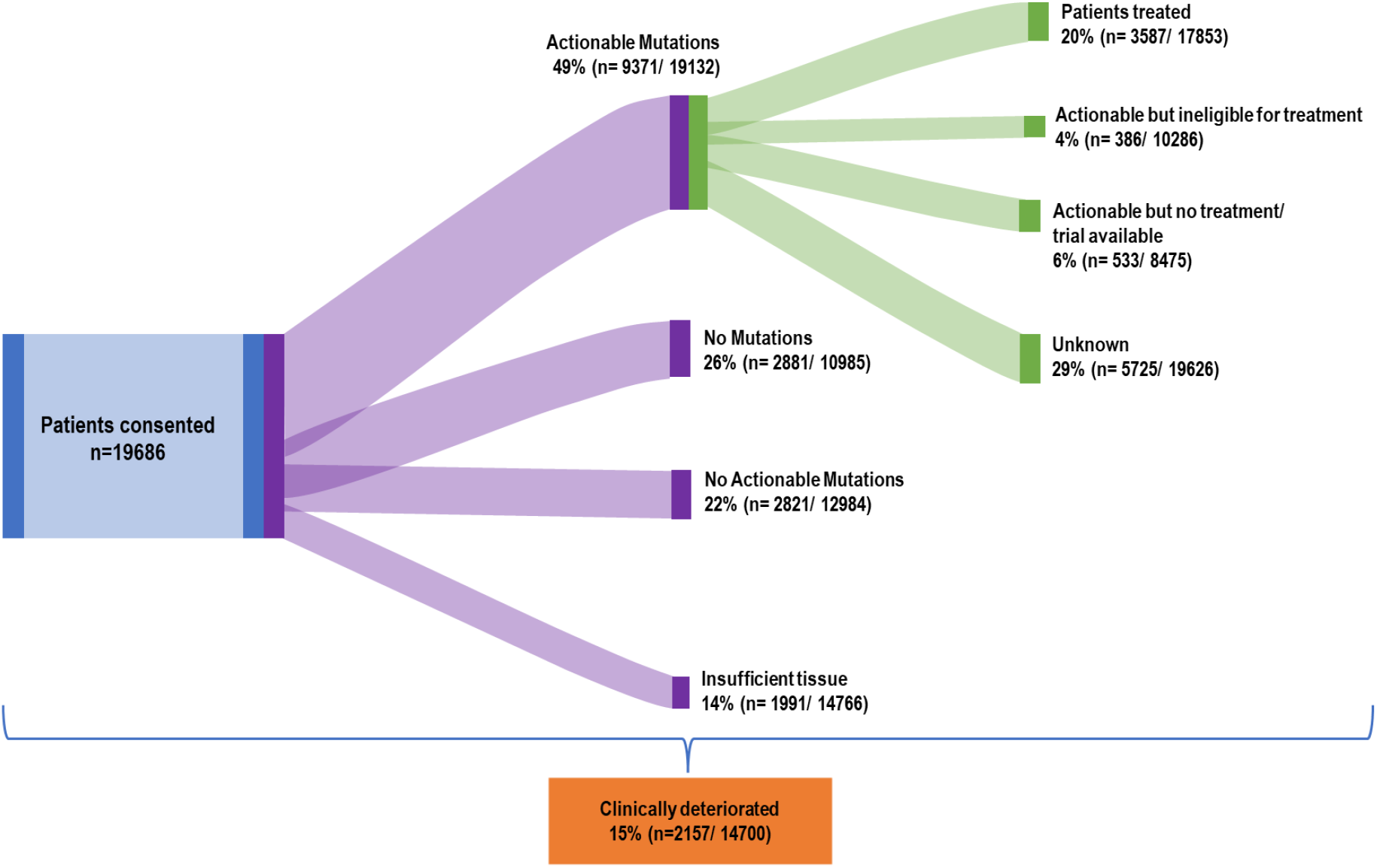
Flow of patients in all studies through an MTB, numbers and percentage are no cumulative as some studies did not report on all reasons for attrition. Clinical deterioration occurred at any stage of the patient journey, and it was not possible to separate these stages out.

In MTBs describing adult patients 20% (3333/16427) went onto treatment; in paediatric studies, 13% (81/628) of patients went on to treatment, and in MTBs where patient populations were both adults and paediatrics 7% (173/2631) went onto treatment. The greatest rates of attrition were due to clinical deterioration, no mutations detected, or no actionable mutations detected.

### Reporting of MTBs

MTBs were inconsistently reported in the literature. Although arguably reasons for patient attrition may vary, some studies failed to report on the number of patients evaluated by an MTB (n=1) and the number of patients treated based on recommendations by MTBs (n=5). Also missing were the composition of the MTB (n=13) and tissue type used for sequencing (n=14). Only 12 MTBs reported on the presence or absence of a cut off for variant allele frequency and 15 reported on what actionability scales were used. Outlined in Figure 1 are the areas in the process where patient attrition occurs and how often this was reported in the literature. Of the data that was available overall 49% of patients had an actionable mutation after genomic profiling and 20% of all patients registered to MTB received recommended treatment. There were no studies that reported on all areas defined in this review.

## Discussion

Molecular tumour boards were developed to assist with assessment of genomic tests to facilitate targeted treatment for patients and have been widely implemented throughout the globe. On average 20% of patients enrolled onto an MTB received an MTB-directed therapy. When able to access treatment, overall response rates vary from 0% - 67% and MTBs at best “do no harm”^66^. MTBs facilitate enrolment of patients on to treatments or trials with biological potential or for their specific tumour type, some may argue that it gives patients the chance, however small, to receive life extending drugs. However, with attrition rates exceedingly high and response rates variable, one patient in 5 will have the opportunity to access a targeted therapy after exhausting standard of care.

As we elicited, there are two key areas of issue with treating patients through an MTB, there are high rates of patient attrition, and secondly there are low response rates. Clinical outcomes are out of the scope of this review, though are covered in detail by Larson et al.^66^ Patient attrition can be summarised into broad categories such as, lack of suitable tissue; no mutations; or actionable mutations but unable to access treatment as not available or; unable to access treatment even though available; and clinical deterioration. Practically addressing specific blockers could facilitate more patients gaining access to treatment, which we address in the following sections.

### Points of Intervention

#### Lack of tissue

All studies in this review, where the sample type used for profiling was specified, used either archived tissue samples, fresh biopsy, or surgical samples as the source of tumour profiling. Where blood samples were taken, this was usually only for germline analysis. Insufficient tissue was one of the most common reasons provided for patient attrition and comprised 14% of all patients (n=1991) in this review. This did not include numbers of patients that were ineligible based upon lack of tissue as they were never enrolled in the MTB. However, to put this into context and highlight the proportion of patients this may omit, in one study alone 3290 patients failed screening due to a lack of tissue and just 229 were enrolled^20^.

Liquid biopsies have been a long awaited tool in oncology. They have been shown to be clinically relevant for different cancer types though there is still much work left to do until they can be utilised routinely by oncologists.^67,68^ However, there is a clear need for alternatives to tissue analysis. Of the four studies that looked at using liquid biopsy for genomic analysis turnaround time data is not available but only 4% (range 0-8%) of patients were ineligible due to a lack of or failure of a sample^12,16,50,55^ compared to 14% (range 0-23%) where analysis failed when only tissue was used (see Supplementary Material table 1). Circulating free tumour derived DNA can be used for the assessment of cancer-specific somatic mutations, chromosomal abnormalities, copy-number alterations and epigenetic modifications and is elevated in malignancy^69^. It has been shown to be useful in therapy selection for patients, particularly in settings where patients are late stage^70,71^. Furthermore, the downsides to tumour biopsies are well documented^72^ and retrieving archived samples can often cause significant delays. Implementing more wide-scale liquid biopsy testing could improve the rates of attrition where patients are lacking sufficient tissue, are unsuitable for biopsy or have old archival samples.

#### No actionable mutations or no mutations

The two greatest reasons for patient attrition within this review were that patients’ sample yielded no mutations or no actionable mutations as determined by the reviewing MTB. It has been shown that more comprehensive multi-omics profiling provides more clinically relevant information^73–75^, however, it is necessary to couple this with technologies that will help to prioritise the inevitable volumes of information produced^76^.

The number of patients accessing treatment changes depending on the types and number of profiling tests that are performed on their sample (e.g. NGS, IHC RNAseq etc.). Interestingly there is an almost 50% decrease in treatment access when two tests are used, and treatment rates do not increase significantly the more complex testing is used (see supplementary material table 2). This could be potentially explained by a lack of drugs associated with these genomic alterations or possibly due to the increased challenge for clinicians at being able to discern potentially actionable mutations due to an excess of data. Developing computational methods to help integrate and interpret data from multiple tests to link with current literature and available treatment options could help to manage the demand on clinicians. A lack of drug availability is also a limiting factor that has the potential to get better over time^77^.

**Table 2.**
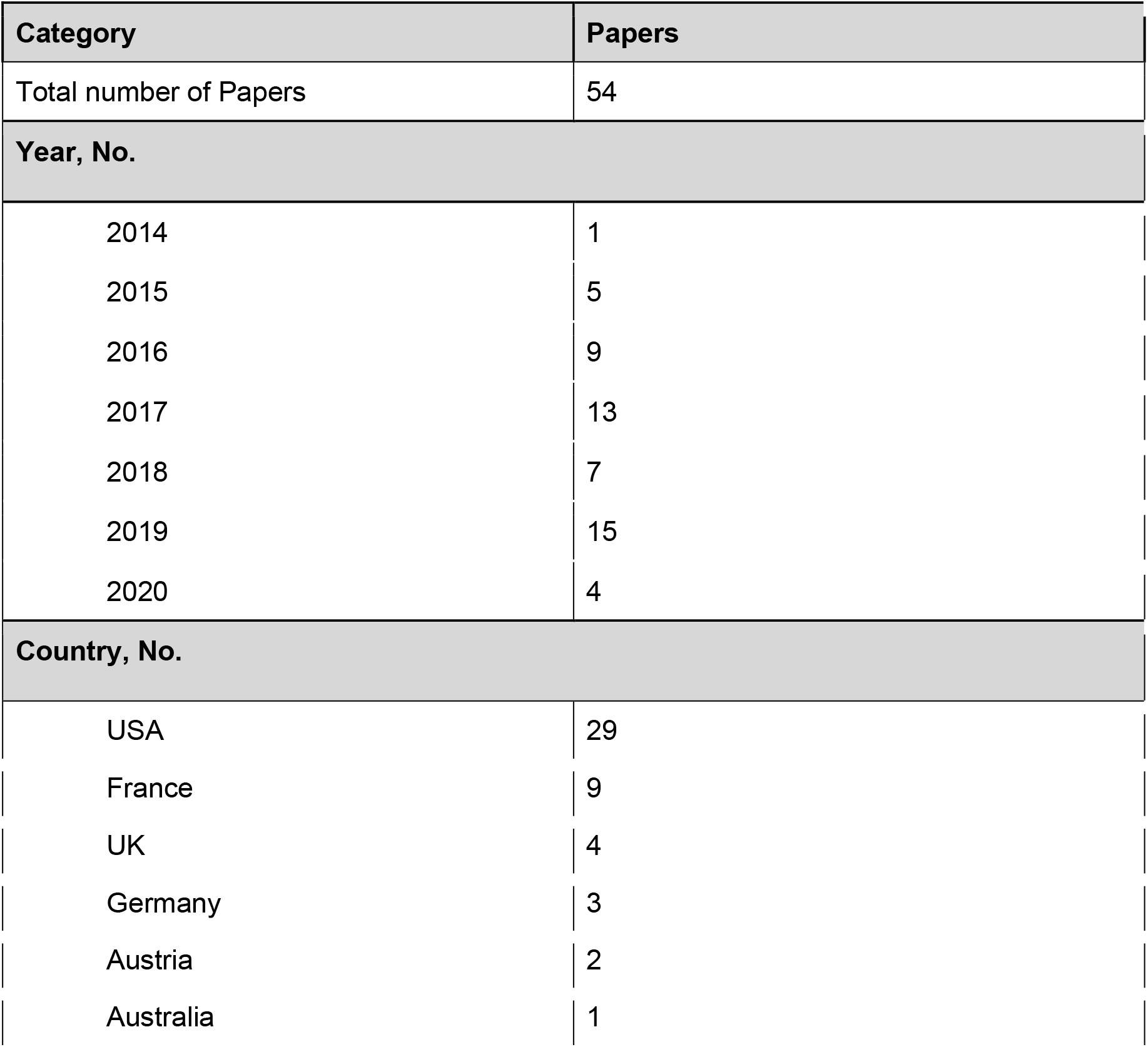
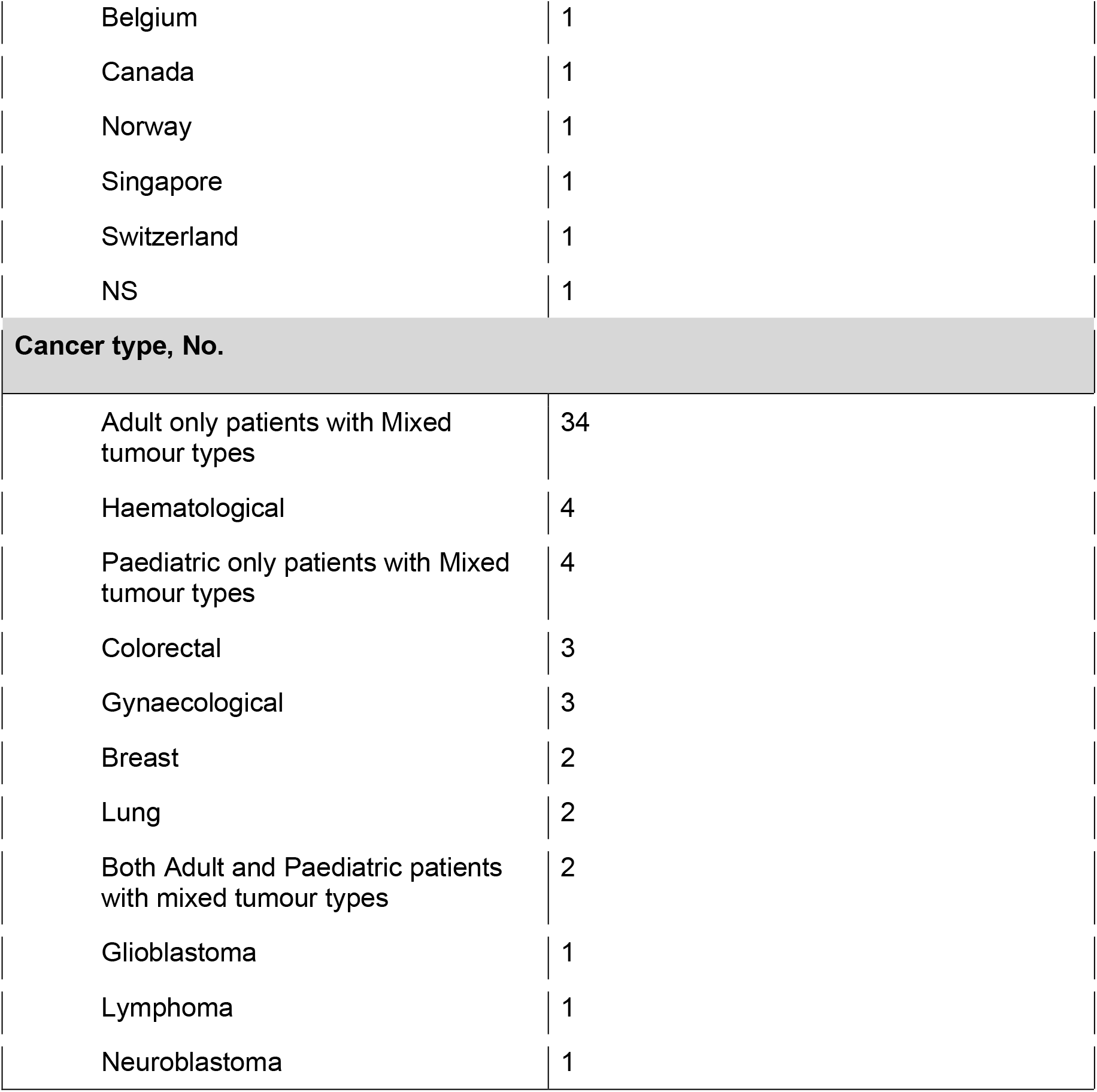
Summary of MTB characteristics

| Category | Papers |
| --- | --- |
| Total number of Papers | 54 |
| Year, No. |  |
| 2014 | 1 |
| 2015 | 5 |
| 2016 | 9 |
| 2017 | 13 |
| 2018 | 7 |
| 2019 | 15 |
| 2020 | 4 |
| Country, No. |  |
| USA | 29 |
| France | 9 |
| UK | 4 |
| Germany | 3 |
| Austria | 2 |
| Australia | 1 |
| Belgium | 1 |
| Canada | 1 |
| Norway | 1 |
| Singapore | 1 |
| Switzerland | 1 |
| NS | 1 |
| **Cancer type, No.** |  |
| Adult only patients with Mixed tumour types | 34 |
| Haematological | 4 |
| Paediatric only patients with Mixed tumour types | 4 |
| Colorectal | 3 |
| Gynaecological | 3 |
| Breast | 2 |
| Lung | 2 |
| Both Adult and Paediatric patients with mixed tumour types | 2 |
| Glioblastoma | 1 |
| Lymphoma | 1 |
| Neuroblastoma | 1 |

**Table 3.**
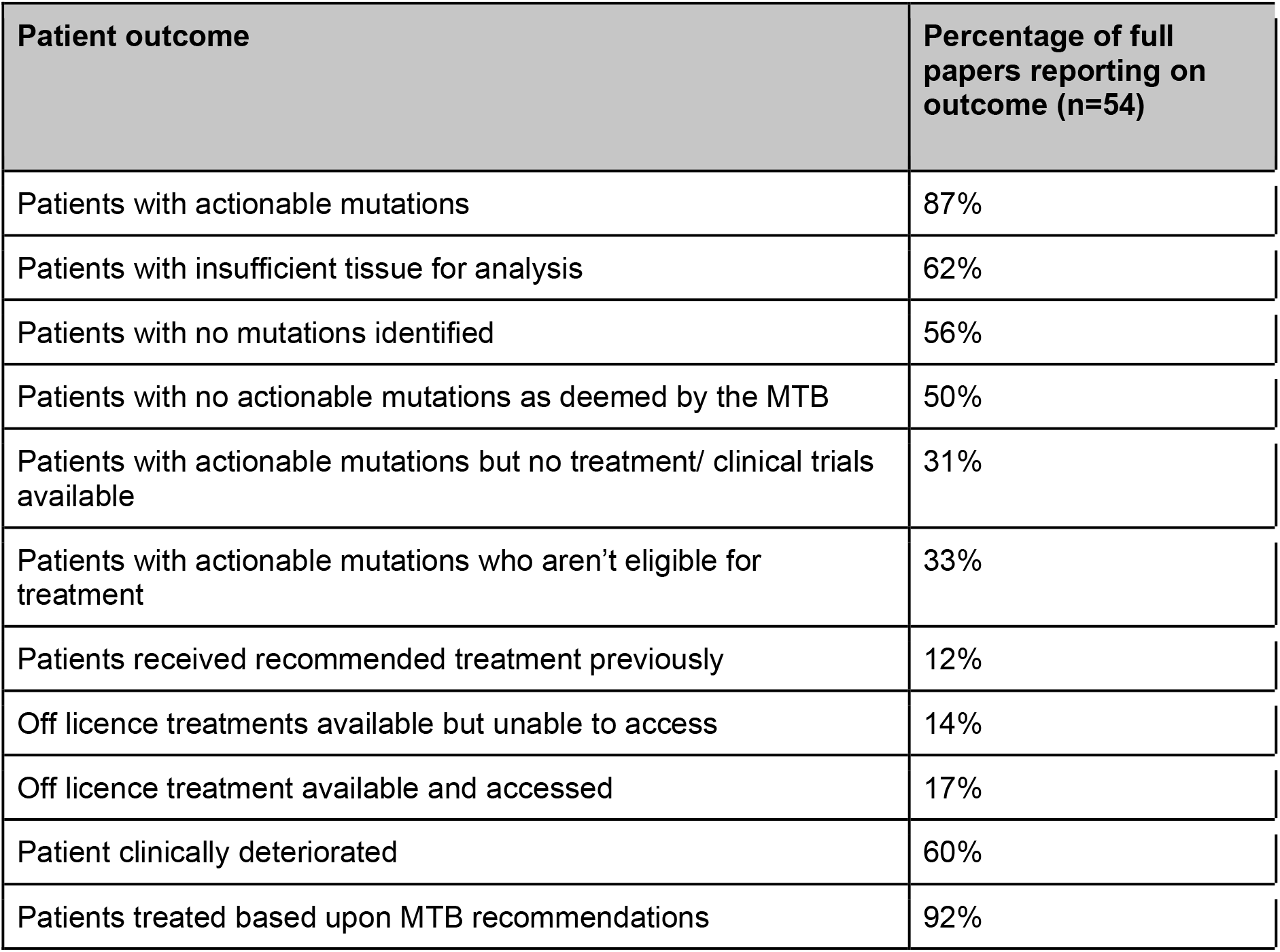
Percentage of papers reporting on specific attrition reasoning within the literature.

#### Actionable mutations but ineligible or no options for treatment

Attrition due to ineligibility for clinical trials or no further treatment options available, both within a clinical trial or using off licence treatment, affected 15% of all patients in this review. However, this was only reported in 13% (off licence treatment available but unable to access) and 31-32% (actionable but no treatment/ trial available or ineligible for treatment) of studies so has the potential to impact greater numbers. Unsurprisingly, a common barrier to paediatric studies was gaining access to treatment options. Though drug development has increased in this area^78^ further work is being done in cancers where presentation in adults differs to children or paediatric-specific cancers^79^.

Eligibility criteria is important to ensure the safety of participants on clinical studies however it can be restrictive, resulting in unjustified exclusion of patients from enrolment into clinical trials^80–83^. A systematic review of randomized controlled trials found that 47.2% of criteria were not scientifically justified^82^. Not only does this result in failed recruitment for studies but also fails to evaluate efficacy and safety in real world populations, and importantly excludes patients from receiving potential treatment options. Therefore, more flexible data-driven eligibility criteria are required to prevent unnecessary exclusion of patients from trials^84,85^, though this will require a wider consensus to drive this change. Importantly, artificial intelligence and machine learning can play a crucial role in evaluating suitable patients for studies that do not follow a restrictive exclusion/ inclusion approach^83^.

Identifying suitable treatment or trial options for patients can be a difficult and onerous task given the large number of recruiting studies, potentially actionable mutations and literature based evidence currently available, which is steadily growing^86^. Therefore, trial matching software can help clinicians review available studies based upon patients profiling results,^87^ not solely relying on clinicians’ knowledge of local and available clinical trials. Some profiling services provide these trial matching services, such as Foundation Medicine^88^, though to date it is unclear how comprehensive or relevant these suggestions are for patients or how often these suggestions are implemented. Additionally, clinical trial slots for dose escalation studies are intermittently available or rapidly fill for small dose escalation cohorts so it is important to be able to capture this rapidly altering data.

Unfortunately, information on accessing off-licence treatment is unavailable, including how often drug applications are accepted or rejected.

#### Patients clinically deteriorated

One of the best reported outcomes was clinical deterioration of patients, whether that was declining performance status, admission to hospice care or death. By the nature of MTBs patients that are considered are often late stage, have rare cancers or poor prognosis. As a result, it is inevitable that they may decline during the process. However, accelerating the process of review will inevitably improve the chances of patients reviewed by MTBs, such as moving to liquid biopsies to reduce wait times for tissue acquisition and preparation or engaging with community teams that refer to the MTBs to facilitate earlier and more accurate referrals, thus decreasing the need for pre-sequencing MTB reviews to evaluate suitability for genomic testing.

### Standardised reporting

In stark contrast to criteria for publishing on clinical trials^89^ there was no standard reporting for MTBs. This is evidenced by the difficulty in obtaining and analysing the data from this review due to the number of missing values. It is important to understand the reasoning behind patient attrition to be able to improve processes and understand barriers to access. Therefore, we suggest a broader community-level discussion on standardised reporting in MTBs. Additionally, providing standard reporting for researchers allow for accurate contrast of approaches, whether that is process or testing driven.

Aiming for a systematic and continuous assessment of attrition and opportunities for therapeutic evolution, we suggest the following categories as a minimum set; tissue type; testing performed; number and types of genetic changes included; variant allele frequency threshold; genomic scale used; number of patients registered to MTBs; number of patients with actionable mutations; attrition numbers and reasoning; total number of patients accessing treatment based on MTB review; and turnaround times from tissue acquisition to discussion at MTB.

## Conclusions

Attrition within MTBs is a pervasive issue that is experienced globally, and as omics-derived data continues to increase alongside new targeted therapies and a growing literature base it is more important than ever to integrate new technologies to guide and aid clinicians in decision making. Consistent reporting is important to understand barriers to accessing treatment via MTBs and more work needs to be done to understand how often patients are unable to access off-licence treatment and clinical trials.

## Data Availability

All data is included in the supplementary materials and via the full papers included in the references.

